# Efficacy and safety of nitazoxanide combined with ritonavir-boosted atazanavir for the treatment of mild to moderate COVID-19

**DOI:** 10.1101/2022.02.03.22270152

**Authors:** Adeola Fowotade, Folasade Bamidele, Boluwatife Egbetola, Adeniyi Francis Fagbamigbe, Babatunde Ayodeji Adeagbo, Bolanle Olufunlola Adefuye, Ajibola Olagunoye, Temitope Olumuyiwa Ojo, Akindele Olupelumi Adebiyi, Omobolanle Ibitayo Olagunju, Olabode Taiwo Ladipo, Abdulafeez Akinloye, Adedeji Onayade, Oluseye Oladotun Bolaji, Steve Rannard, Christian Happi, Andrew Owen, Adeniyi Olagunju

**Affiliations:** Department of Medical Microbiology and Parasitology, University of Ibadan, Ibadan, Nigeria; Olabisi Onabanjo University Teaching Hospital, Sagamu, Nigeria; Department of Epidemiology and Medical Statistics, University of Ibadan, Ibadan, Nigeria; Department of Pharmaceutical Chemistry, Obafemi Awolowo University, Ile-Ife, Nigeria; State Specialist Hospital, Osogbo, Nigeria; Department of Community Health, Obafemi Awolowo University Teaching Hospital, Ile-Ife, Nigeria; Department of Community Medicine, University of Ibadan, Ibadan, Nigeria; Department of Surveillance and Epidemiology, Nigeria Centre for Disease Control, Abuja, Nigeria; Oyo State Ministry of Health, Ibadan, Nigeria; Department of Chemistry, University of Liverpool, United Kingdom; African Centre of Excellence for Genomics of Infectious Diseases, Redeemer’s University, Ede, Nigeria; Department of Pharmacology and Therapeutics, University of Liverpool, Liverpool, United Kingdom

## Abstract

**Background:** Finding effective therapeutics for COVID-19 continues to be an urgent need, especially considering use context limitations and high cost of currently approved agents. The NACOVID trial investigated the efficacy and safety of repurposed antiprotozoal and antiretroviral drugs, nitazoxanide and atazanavir/ritonavir, used in combination for COVID-19.

**Methods:** In this pilot, randomized, open-label trial conducted in Nigeria, patients diagnosed with mild to moderate COVID-19 were randomly assigned to receive standard of care (SoC) or SoC plus a 14-day course of nitazoxanide (1000 mg b.i.d.) and atazanavir/ritonavir (300/100 mg od) and followed through day 28. Study endpoints included time to clinical improvement, SARS-CoV-2 viral load change, and time to complete symptom resolution. Safety and pharmacokinetics of nitazoxanide active metabolite, tizoxanide, were also evaluated. This trial was registered with ClinicalTrials.gov (NCT04459286).

**Findings:** There was no difference in time to clinical improvement between the SoC (n = 26) and SoC plus intervention arms (n = 31; Cox proportional hazards regression analysis adjusted hazard ratio, aHR = 0.898, 95% CI: 0.492-1.638, p = 0.725). No difference was observed in the pattern of saliva SARS-CoV-2 viral load changes from days 2 to 28 in the 35% of patients with detectable virus at baseline (20/57) between the two arms (aHR = 0.948, 95% CI: 0.341-2.636, p = 0.919). There was no significant difference in time from enrolment to complete symptom resolution (aHR = 0.535, 95% CI: 0.251 - 1.140, p = 0.105). Atazanavir/ritonavir increased tizoxanide plasma exposure by 68% and median trough plasma concentration was 1546 ng/ml (95% CI: 797-2557), above its putative EC_90_ in 54% of patients. Tizoxanide was not detectable in saliva.

**Interpretation:** These findings should be interpreted in the context of incomplete enrolment (64%) and the limited number of patients with detectable SARS-CoV-2 in saliva at baseline in this trial.

**Funding:** The University of Liverpool.

**Research in context:** *Evidence before this study:* The potential efficacy of nitazoxanide as a repurposed drug for COVID-19 is being investigated in a number of studies due to confirmed *in vitro* activity against SARS-CoV-2. Available data from completed randomised controlled trials in which clinical improvement, effect on viral load, and symptom resolution were evaluated as outcomes do not offer conclusive evidence.

*Added value of this study:* In the NACOVID trial, we sought to take advantage of a model-informed strategy and known interaction between nitazoxanide and atazanavir/ritonavir to achieve optimal concentration of tizoxanide in plasma, and possibly in respiratory tracts of patients with mild to moderate COVID-19. While this strategy significantly enhanced tizoxanide exposure in the plasma of patients, our data indicated poor penetration into the respiratory tracts. Specifically, there were no differences in time to clinical improvement, viral load changes, and symptom resolutions between patients who were given standard of care alone and those who combined it with study intervention.

*Implications of all the available evidence:* The clinical benefit of nitazoxanide remains uncertain. The present study highlights the need for early insight into target site biodistribution of potential COVID-19 therapeutics to better inform candidate selection for clinical trials.

## Introduction

With over 350 million cases and more than 5.6 million deaths at the end of January 2022,^1^ just over 24 months since the first case was reported in mainland China,^2^ the coronavirus disease 2019 (COVID-19) is by far the most devastating pandemic known to anyone alive.^3^ More than 2900 vaccine or therapeutic clinical trials have been registered and hundreds are either completed or ongoing.^4^ Remdesivir, an intravenous nucleotide prodrug originally developed for Ebola virus disease,^5^ was the first drug approved by the FDA in October 2020. Emergency use authorisations were issued in December 2021 for paxlovid^6^ and molnupiravir,^7^ both orally administered antiviral agents. Though non-pharmaceutical interventions have helped to break the chain of transmission and global deployment of effective vaccines has reduced disease severity, there is an urgent need for additional effective therapeutics for treatment and/or prevention of COVID-19.

In a report of *in vitro* studies on the anti-coronavirus activity of 727 compounds in the National Institutes of Health Clinical Collection small molecule library, 84 drugs with significant anti-coronavirus activity were identified, including 51 entry blockers and 19 inhibitors of viral replication in cell culture using a luciferase reporter-expressing recombinant murine coronavirus.^8^ Nitazoxanide was among the top three inhibitors, resulting in a reduction of 6 log_10_ in virus titre with an IC_50_ of 1.0 µM. The major circulating metabolite of nitazoxanide is tizoxanide and recent work by the NIH National Centre for Advancing Translational Sciences confirmed its *in vitro* activity against SARS-CoV-2 in Vero E6 host cells *via* suppression of viral cytopathic effect.^9^ We previously identified nitazoxanide among the only 14 drugs able to achieve plasma and lung concentration above the EC_90_ for SARS-CoV-2 at approved doses out of 56 drugs with reported *in vitro* activity.^10^ In a follow-up study, we explored optimal nitazoxanide dosing schedules for maintaining effective tizoxanide plasma and lung concentrations.^11^ The susceptibility of 210 seasonal influenza viruses to nitazoxanide and its metabolite tizoxanide has been reported^12^ and nitazoxanide reduced symptom duration in acute uncomplicated influenza^13^. SARS-CoV-2 shares almost 80% of the genome with SARS-CoV^14^ and almost all encoded proteins of SARS-CoV-2 are homologous to SARS-CoV proteins.^15^ Hence, nitazoxanide and its metabolite tizoxanide with demonstrated *in vitro* activity against SARS-CoV are considered potential candidates for COVID-19. The HIV protease inhibitor, atazanavir (boosted with ritonavir), has been shown to inhibit the major protease enzyme required for viral polyprotein processing during coronavirus replication.^16,17^ It also blocks pro-inflammatory cytokine production.^16^ Additionally, tizoxanide is inactivated by glucuronidation and atazanavir is a well-known inhibitor.^18^ Hence, atazanavir is expected to enhance tizoxanide exposure when used in combination with nitazoxanide. Drug repurposing often requires consideration of target concentrations and dosing regimens that may not be identical to previously defined labels where optimization was conducted for a different disease. Importantly, widespread deployment of antiviral monotherapies for pulmonary viruses (e.g. influenza virus) often leads to the emergence of resistance and we previously called for caution in this regard.^19^ Therefore, to take advantage of the anticipated favourable drug-drug interaction, a combination of nitazoxanide and atazanavir/ritonavir was selected for this trial.

## Methods

### Study design

The nitazoxanide plus atazanavir/ritonavir for COVID-19 (NACOVID) trial is a pilot open-label randomised phase 2, multicentre, two-arm controlled trial conducted in Nigeria. Patients who recently tested positive for SARS-CoV-2 by means of reverse transcription-polymerase chain reaction (RT-PCR) assay and were symptomatic were eligible. Patients were considered to have a mild disease if they were ambulatory, need little or no assistance. Those with moderate disease were non-ambulatory but had no need for oxygen therapy, or required oxygen by mask or nasal prongs. Severely ill patients that required high-flow nasal cannula oxygen or mechanical ventilation at screening, or had sepsis with end-organ involvement were not eligible. The national guideline for COVID-19 at the time required that all symptomatic cases be managed in isolation and treatment centres established within tertiary hospitals or purpose-built facilities. Hence, the NACOVID trial was conducted in an inpatient setting with participants enrolled after diagnosis and within 48 h of admission.

The National Health Research Ethics Committee, Nigeria (approval number: NHREC/01/01/2007-26/08/2020) and the Central University Research Ethics Committee, University of Liverpool (reference number: 8074) approved the protocol. The National Agency for Food and Drug Administration and Control in Nigeria authorised the trial and independent oversight was provided by a Data and Safety Monitoring Board (DSMB) that included five members with expertise in infectious diseases, clinical trials, pharmacology, and public health. A medical monitor conducted independent monitoring visits to trial sites in line with the Clinical Trial Monitoring Plan to ensure the safety of participants and compliance with approved protocol. All patients provided written informed consent as per the ethics committee’s approved process. Further details about the trial design, inclusion and exclusion criteria are provided in the published protocol.^20^ The trial is registered on ClinicalTrials.gov (NCT04459286) and Pan African Clinical Trials Registry (PACTR202008855701534).

### Randomisation

Patients were randomly assigned in a 1:1 ratio to receive either standard of care alone or standard of care combined with 1000 mg nitazoxanide tablets twice daily and 300/100 mg atazanavir/ritonavir tablets once daily. Randomisation was implemented using a Research Electronic Data Capture (REDCap)^21^ module that centrally stratified patients by study site, diagnosis CT value, gender, existence of comorbidities and disease severity at enrolment. Standard of care was according to the national interim guidelines for clinical management of COVID-19, including antipyretics for fever, cough medicine, antimalaria in cases with malaria co-infection, multivitamins and mineral supplement, and ongoing treatment of pre-existing comorbidities. Participation in other interventional studies or off-label use of other medications intended as specific treatment for COVID-19 outside the standard of care was not allowed throughout the 28-day study period.

### Procedures

On study day 0 (baseline), patients provided informed consent and were assessed for eligibility. Site clinical investigators documented demographic and anthropometric information, recent and current medical history including confirmation of COVID-19 diagnosis and disease severity, pregnancy test for reproductive age women, concomitant medications, physical examination, vital signs, and safety blood for haematology and biochemistry. Those who met the eligibility criteria were enrolled and randomised to either continue the standard of care alone (started before study entry in all participants) or trial intervention in addition. The intervention consisted of 1000 mg nitazoxanide twice daily and 300/100 mg atazanavir/ritonavir administered orally once daily in the night, both administered orally after a meal and directly observed by study staff on days 1 to 14.

Daily assessment of vitals including SpO_2_, symptom monitoring using the Flu-PRO questionnaire and clinical improvement as well as adverse event monitoring was conducted by designated staff at each study site for all patients on days 1 to day 14, and on days 21 and 28. Saliva for SARS-CoV-2 viral load was collected on days 0, 2, 4, 6, 7, 14, and 28. Saliva and dried blood spots for quantification of tizoxanide, the active metabolite of nitazoxanide, were collected on days 2, 4, 6, 7, and 14 about the same time as viral load samples. Patients who were discharged from the isolation and treatment centre after 14 days in line with the national guideline for clinical management of COVID-19 returned to site for days 21 and 28 follow-up. All samples were stored on-site at -20°C, or lower, and shipped to the testing laboratories: SARS-CoV-2 viral load at the African Centre of Excellence in Genomics of Infectious Diseases (ACEGID), Redeemers University, Ede and pharmacokinetic analysis at the Bioanalytical Laboratory, Obafemi Awolowo University, Ile-Ife, Nigeria. Study data were collected and managed with a 26-form electronic case report form using REDCap,^21^ a secure, web-based software platform designed to support data capture for research studies hosted at Obafemi Awolowo University.

### Outcomes

The main outcomes were time to SARS-CoV-2 RT-PCR negativity, time to clinical improvement, and temporal patterns of saliva SARS-CoV-2 viral load quantified by RT-PCR. Clinical improvement was defined as the time from randomisation to either an improvement of two points on a 10-category ordinal scale or discharge from the hospital, whichever came first. The ordinal scale was developed by the WHO Working Group on the Clinical Characterisation and Management of COVID-19 infection^22^ with the following categories: 0, uninfected with no viral RNA detected; 1, asymptomatic with viral RNA detected; 2, symptomatic but independent; 3, symptomatic and in need of assistance; 4, hospitalized but not requiring oxygen therapy; 5, hospitalised and requires oxygen by mask or nasal prongs; 6, hospitalized and requires oxygen by NIV or high flow; 7, intubated and on mechanical ventilation with PaO2/FiO2 ≥150 or SpO2/FiO2 ≥200; 8, on mechanical ventilation with PaO2/FiO2 ˂150 (SpO2/FiO2 ˂200) or vasopressors; 9, on mechanical ventilation with PaO2/FiO2 ˂150 and vasopressors, dialysis, or extracorporeal membrane oxygenation; 10, dead. Secondary outcomes included time to symptom resolution, clinical status on days 7 and 14 based on the 10-category ordinal scale, day 28 mortality, time from treatment initiation to death and proportion of participants with viral RNA detection over time.

For the assessment of pharmacokinetic endpoints, sparse dried blood spots samples were collected on Whatman 903 protein saver cards (VWR International Ltd, Leicestershire, UK) at steady state to determine the mid-dose concentration of tizoxanide, the active metabolite of nitazoxanide. Saliva samples for tizoxanide quantification were collected at the same time as saliva samples for SARS-CoV-2 viral load on days 2, 4, 6, 7 and 14. The drug-drug interaction potential of nitazoxanide and atazanavir/ritonavir was investigated in a separate healthy volunteer, two-period cross-over study. In brief, drug-free healthy volunteers (18-35 years old, male and female) were recruited. Each volunteer received 1000 mg nitazoxanide 12 hourly after a standard meal for 5 days in the first period, followed by a 21-day washout period. In the second stage, they received 1000 mg nitazoxanide 12 hourly combined with 300/100 mg atazanavir/ritonavir once daily for 5 days. Plasma samples were collected at 0.25, 0.5, 1, 2, 4, 6, and 12 hours after dose on days 1 and 5 during both stages. Tizoxanide quantification was by validated LC-MS/MS methods on TSQ Vantage (Thermo Electron Corporation, Hemel Hempstead, Hertfordshire, UK) with 50 ng/ml lowest limit of quantification. Data from the first seven participants who completed day 1 of both periods are included in this paper to show the outcome of single-dose interaction. The full study, including an embedded clinical cross validation of the plasma and dried blood spot bioanalytical methods, will be published separately.

### Statistical analysis

A sample size of 98 was estimated to provide more than 80% power to show or exclude 60% improvement in time to SARS-CoV-2 RT-PCR negativity in the intervention group compared with the control group at a two-sided type 1 error rate of 5%. Between-group (SOC vs Intervention) comparisons of demographic, anthropometric, clinical and laboratory data of the participants were conducted using independent sample t-test and Chi-square test of association for continuous and categorical variables respectively. Analysis of clinical improvement based on the 10-category ordinal scale was performed using the analysis of time-to-event data. Multivariable Cox proportional hazard regression analysis was conducted to assess the differentials in time to improvement. Analysis of cumulative (probability of survival) improvement rate was carried out using Kaplan-Meier survival curves. Primary and secondary outcomes analyses were adjusted for the baseline value of the outcome and randomization stratification factors. SARS-CoV-2 viral load was calculated from the RT-PCR cycle-threshold value. Daily symptom data were aggregated per category (nose and throat, eyes, chest and respiratory, gastrointestinal, and body and systemic) and complete resolution was defined as the disappearance of all abnormalities. Covariates with p value < 0.25 in the univariable Cox regression analysis were included in the multivariable model. These analyses were conducted using Stata ® Version 17.

### Role of funding source

There was no external funding source for this study.

## Results

The first patient was enrolled on November 25, 2020 and the last patient was enrolled on April 20, 2021. To take advantage of the increasing cases during the second wave of the pandemic in Nigeria, two under-recruiting sites (Obafemi Awolowo University Teaching Hospitals Complex, Ile-Ife and State Specialist Hospital, Osogbo) were withdrawn on February 1, 2021 and a new site (ThisDay Dome COVID-19 Isolation and Treatment Centre, Abuja) was added. However, no patient was enrolled from the new site as the second wave entered the decline phase before ethics and regulatory approvals of the amendments were secured. Hence, only 57 patients were successfully enrolled and randomised from the Infectious Diseases Hospital, Olodo, Ibadan (n = 45) and Olabisi Onabanjo University Teaching Hospital, Sagamu (n = 12). A total of 26 patients were randomised to the standard of care alone arm and 31 were randomised to the standard of care plus intervention arm (Figure 1). Four patients who complained about the size of the intervention tablets and requested to stop (two on day 2 and two on day 4) were retained in the standard of care alone arm of the trial. A fifth patient in the intervention arm who requested transfer to the standard of care alone arm after 4 days for no clear reason was withdrawn by site investigators. All available data were used in the analysis; excluding patients who switched arms or were either withdrawn gave similar results. Hence, withdrawn participant data were censored as of the withdrawal date, while those who switched arms were censored as of the day of switching.

**Figure 1.**
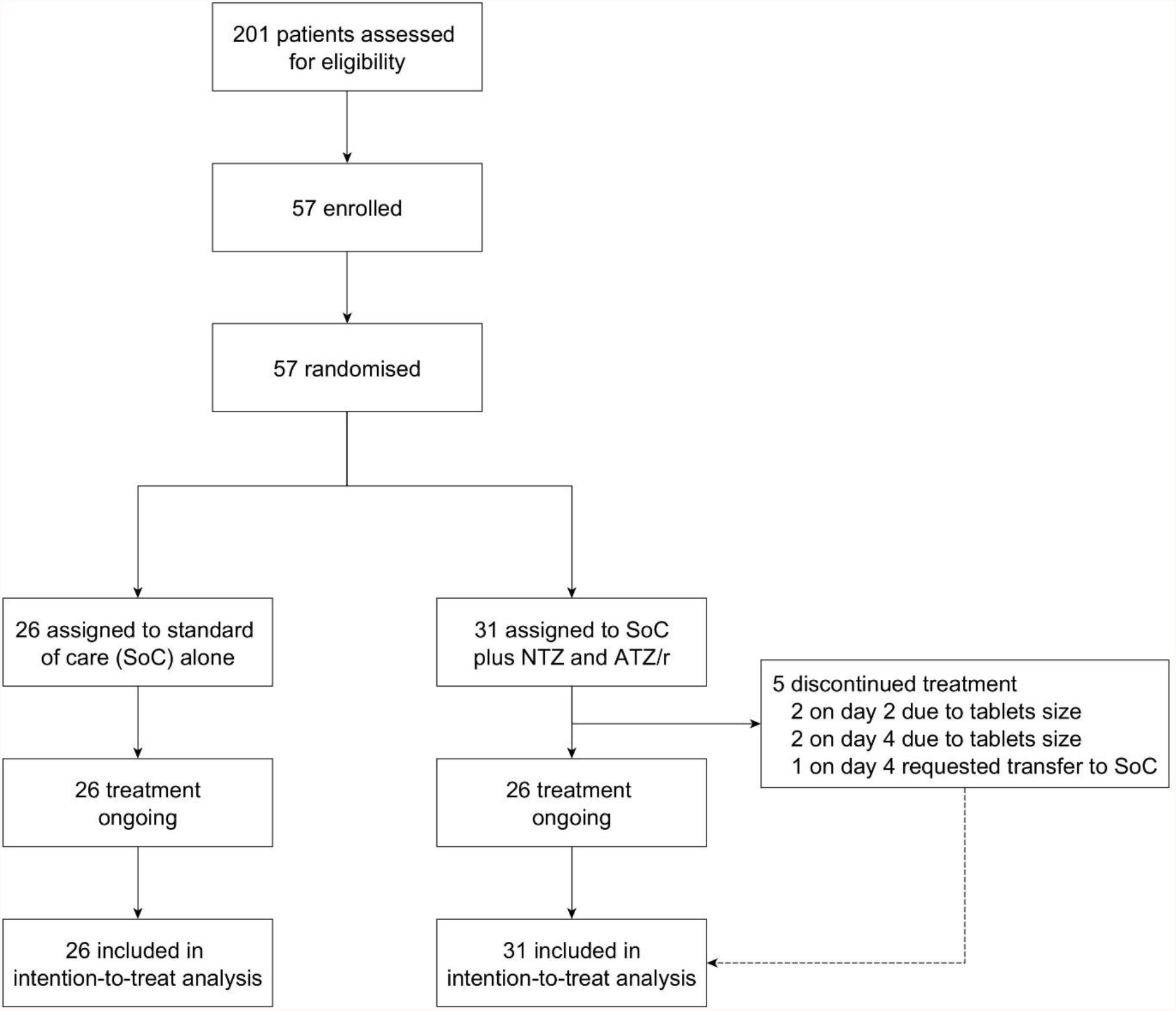
NACOVID trial profile.

### Baseline characteristics

The mean age of patients in the standard of care alone arm was 40 years (standard deviation: 18) and 37 years (13) in the standard of care plus intervention arm. Most participants were male with mean body weights of 67 kg and 70 kg, respectively. In both arms, about 50% of patients were enrolled within 1-4 days of receiving their diagnosis. All baseline characteristics were similar between both groups (Table 1).

**Table 1.** Baseline characteristics of NACOVID trial participants at enrolment.

| Participants | All<br>(N = 57) | SoC alone<br>(N = 26) | SoC with NTZ/<br>ATZ/r (N = 31) | P value |
| --- | --- | --- | --- | --- |
| **Body weight (kg)** | 68 (11) | 67 (11) | 70 (11) | 0.322 |
| **Body mass index (kg/m<sup>2</sup>)** |  |  |  |  |
| Underweight (<18.5) | 3 (5) | 2 (8) | 1 (3) | 0.397 |
| Normal weight (18.5-24.9) | 21 (37) | 11 (42) | 10 (32) |  |
| Overweight (24.5-29.9) | 22 (39) | 7 (27) | 15 (48) |  |
| Obese (≥30) | 11 (19) | 6 (23) | 5 (16) |  |
| **Age (years)** | 38 (16) | 40 (18) | 37 (13) | 0.620 |
| 18-50 | 37 (65) | 18 (69) | 19 (61) | 0.532 |
| 51-75 | 20 (35) | 8 (31) | 12 (39) |  |
| **Sex** |  |  |  |  |
| Female | 19 (33) | 7 (27) | 12 (39) | 0.347 |
| Male | 38 (67) | 19 (73) | 19 (61) |  |
| **Ethnicity** |  |  |  |  |
| Hausa | 2 (4) | 1 (4) | 1 (3) | 0.921 |
| Igbo | 3 (5) | 1 (4) | 2 (7) |  |
| Yoruba | 44 (77) | 21 (81) | 23 (74) |  |
| Others | 8 (14) | 3 (12) | 5 (16) |  |
| **Comorbidities** |  |  |  |  |
| No | 42 (74) | 16 (62) | 26 (84) | 0.057 |
| Yes | 15 (26) | 10 (39) | 5 (16) |  |
| **Time from diagnosis to enrolment (days)** |  |  |  |  |
| ≤ 1 days | 10 (18) | 13 (50) | 15 (48) | 0.468 |
| 2-4 days | 29 (51) | 7 (27) | 5 (16) |  |
| ≥ 5 days | 18 (31) | 6 (23) | 11 (36) |  |
| **Disease severity** |  |  |  |  |
| Mild Covid-19 | 44 (77) | 19 (73) | 25 (81) | 0.571 |
| Moderate Covid-19 | 10 (18) | 6 (23) | 4 (13) |  |
| Severe Covid-19 | 3 (5) | 1 (4) | 2 (6) |  |
| **Baseline symptoms** |  |  |  |  |
| Nose and throat | 57 (100) | 26 (100) | 31 (100) | 1.000 |
| Chest/respiratory | 21 (37) | 10 (39) | 11 (35) | 0.816 |
| Gastrointestinal | 3 (5) | 1 (4) | 2 (7) | 0.661 |
| Body/systemic | 15 (26) | 7 (27) | 8 (26) | 0.924 |
| **Ct value at diagnosis** | 28.4 (6.9) | 29.7 (11.2) | 29.3 (6.9) | 0.338 |
| **Saliva SARS-CoV-2 viral load (copies/ml)** | 127094<br>(337070) | 112256<br>(325927) | 149352<br>(374849) | 0.546 |
| **SPO<sub>2</sub> %** | 97.9 (4) | 97.5 (0.64) | 98.3 (0.37) | 0.263 |

Ct, cycle threshold on the reverse transcriptase polymerase chain reaction assay; SARS-CoV-2, severe acute respiratory syndrome coronavirus 2; SPO<sub>2</sub>, peripheral oxygen saturation.

### Primary outcomes

At the time of enrolment, 19 of the 26 patients randomised to the standard of care alone arm were graded 5 (required oxygen by mask or nasal prongs), 3 were graded 4 (non-ambulatory but did not require oxygen therapy), 1 was graded 2 (symptomatic but ambulatory and independent), and 3 were graded 1 (asymptomatic and ambulatory). Of the 31 patients randomised to the standard of care plus intervention, 23 were graded 5, 3 were graded 4, 2 were graded 3 (ambulatory, but symptomatic and need assistance), 1 was graded 2, and 2 were graded 1. The time to achieve protocol-defined clinical improvement (a drop of 2 levels on the 1-10 ordinal scale) in the entire cohort was 7 days and no difference was observed between the two arms (7 days in the standard of care arm alone versus 8 days in the standard of care plus intervention arm). The hazard ratio (HR) was 1.027 (95% CI: 0.592-1.783), p = 0.924 and no difference was observed after adjusting for potential co-founders in Cox proportional hazards regression analysis, including randomisation stratification variables (aHR = 0.898, 95% CI: 0.492-1.638, p = 0.725; Figure 2). In a separate analysis, we further explored time to clinical improvement in various subgroups using logrank tests but found no significant differences between both arms (Table S1).

**Figure 2.**
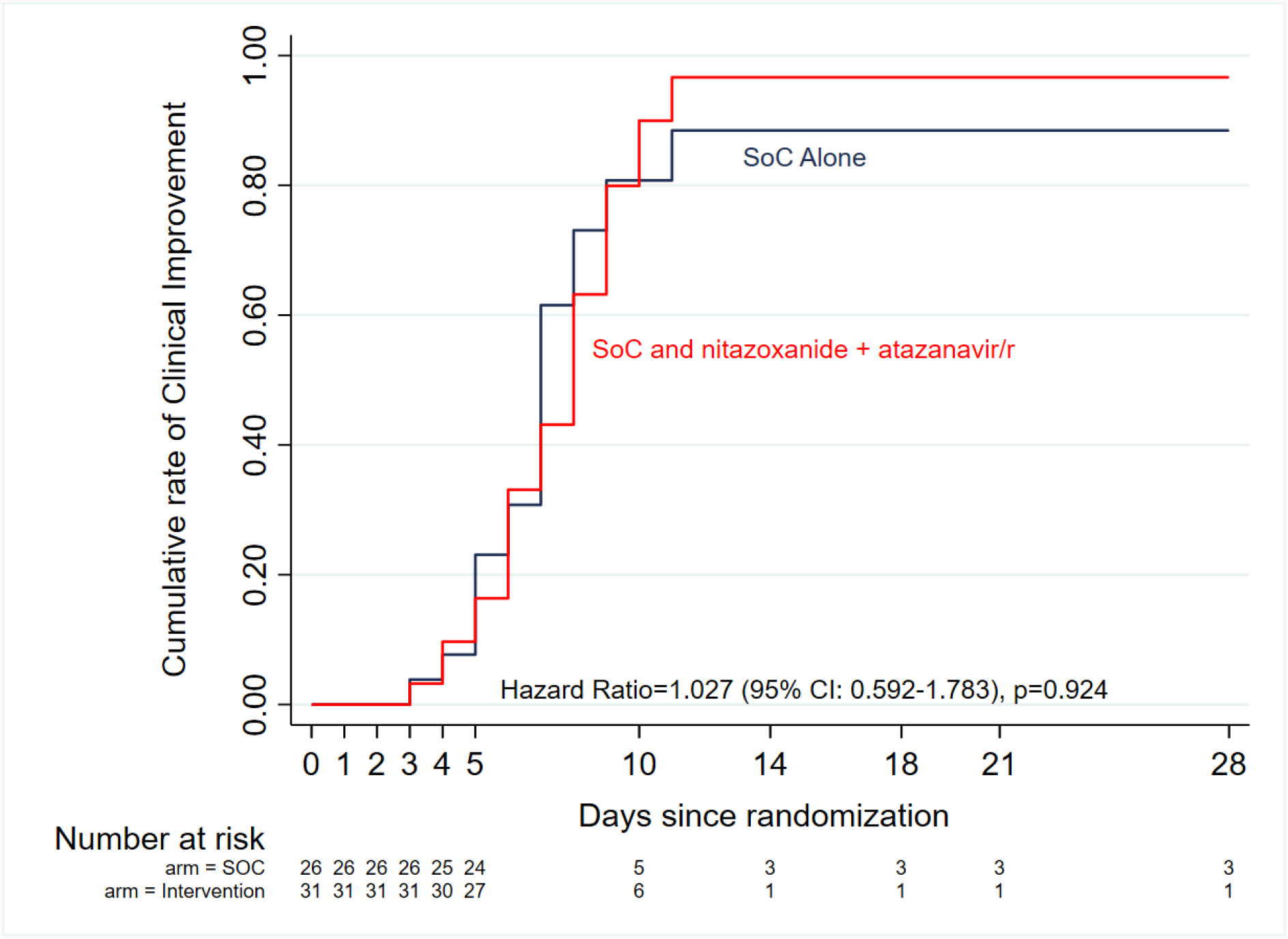
Kaplan–Meier curves of time to clinical improvement (defined as a drop of 2 levels on the 1-10 ordinal scale) by study arm. There was no difference between the two arms (7 days in the standard of care arm alone versus 8 days in the standard of care plus intervention arm). The Cox proportional hazards model adjusted hazard ratio was 0.898 (95% CI: 0.492-1.638, p = 0.725) after adjusting for potential co-founders, including randomisation stratification variables, age and sex.

SARS-CoV-2 was detectable in saliva samples collected at enrolment only in 35% (20/57) of patients with a mean of 5.05 log_10_ copies/ml in SoC alone arm, and 5.17 log_10_ copies/ml in SoC plus intervention arm. In a very limited analysis of this outcome using days 2, 4, 6, 7, 14 and 28 follow up saliva viral load data from these patients, there was no trend towards a difference in the pattern of viral load changes between the two arms, Welch’s t-test p value = 0.758 for comparison of means over the follow-up period (Figure 3). The aHR was 0.948 (0.341-2.636) with a p value of 0.919.

**Figure 3.**
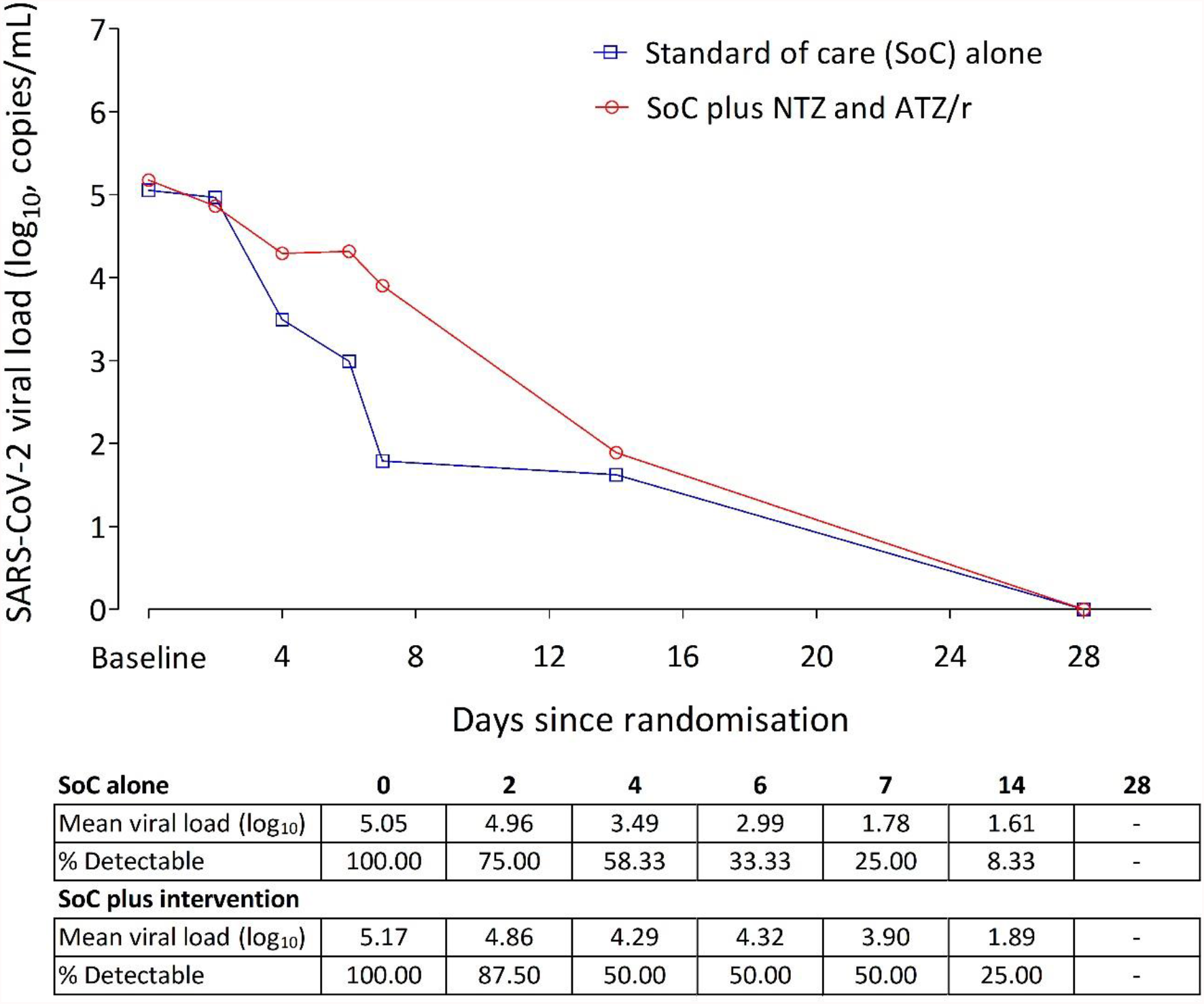
Changes in SARS-CoV-2 viral load in saliva of patients from enrolment to study day 28. In the 20 patients with detectable saliva viral load at enrolment, baseline viral load was 5.05 log10 copies/ml in the SoC alone arm (n = 12), and 5.17 log10 copies/ml in the SoC plus intervention arm (n = 8). In this small cohort, there was no difference in the rate of viral load decline between the two arms (Cox proportional hazards model aHR = 0.948, 95% CI: 0.341-2.636, p = 0.919).

### Secondary and safety outcomes

The median time from enrolment to complete symptom resolution was 8 days in the entire cohort, with a non-significant trend (Kaplan Meier HR = 0.617 (95% CI: 0.311-1.224, p = 0.167) towards a shorter time in the standard of care alone arm (6 days) compared with standard of care plus intervention arm (10 days) (Figure 4). Multivariable Cox proportional hazards regression analysis adjusting for randomisation variables showed a similar overall non-significant trend (aHR = 0.535, 95% CI: 0.251 -1.140, p = 0.105), except for disease severity where moderately ill patients were 67% more likely to achieve complete symptom resolution if they received standard of care alone compared with standard of care plus intervention (aHR = 0.322 (95% CI: 0.122-0.848, p = 0.022). Further exploration of median time to complete symptom resolution in various subgroups using logrank tests showed no trend towards any benefit in combining the intervention with the standard of care (Table S2).

**Figure 4.**
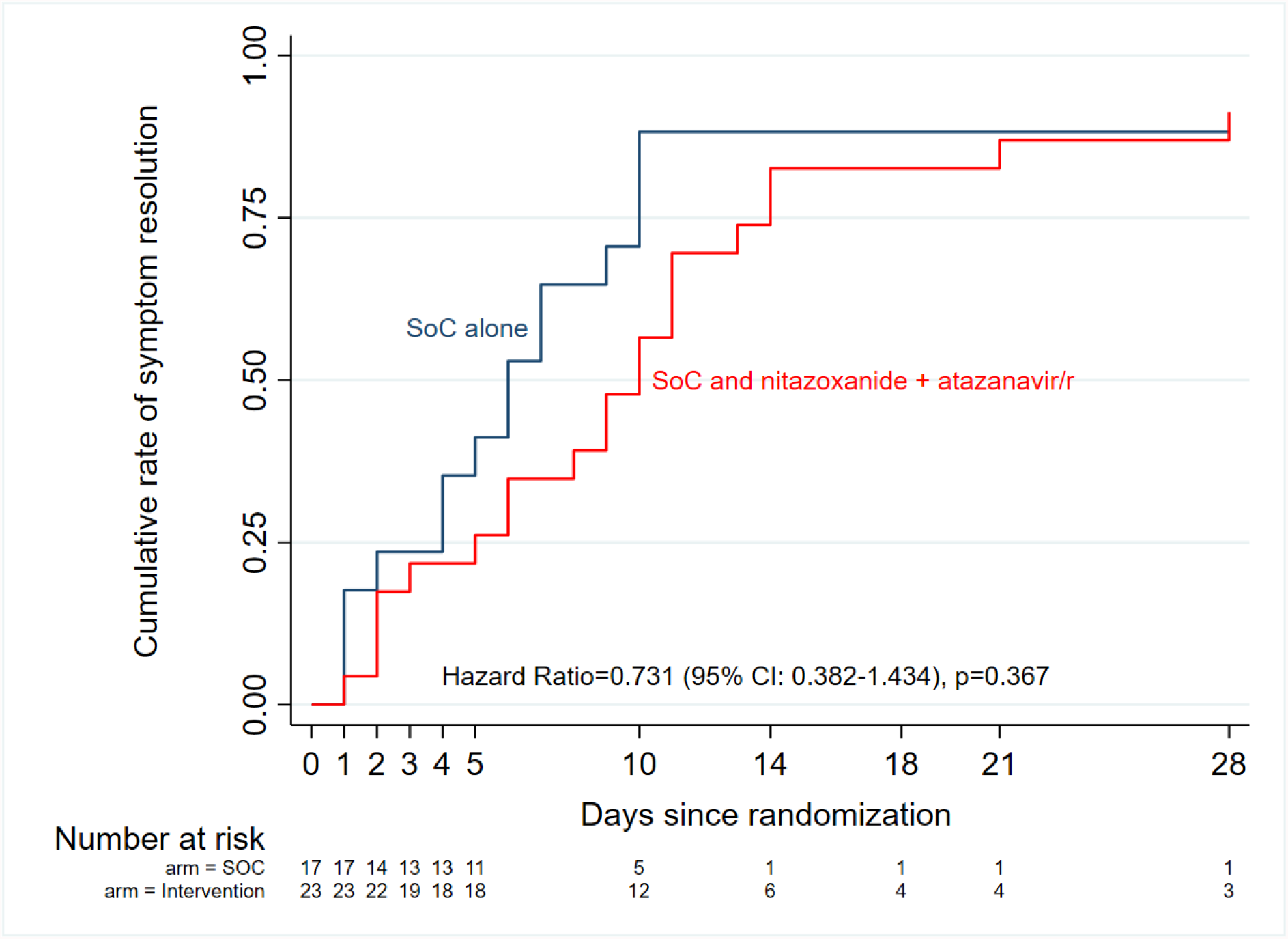
Kaplan–Meier curves of median time to complete symptom resolution by study arm. Overall, there was no significant difference between the two arms, even after adjusting for potential co-founders (Cox proportional hazards model aHR = 0.535, 95% CI: 0.251 -1.140, p = 0.105).

The DSMB at their meeting of 14 November 2021 recommended terminating the trial as no further opportunities existed to recruit additional patients and accrued data did not indicate any trend of benefit in adding the intervention to the standard of care.

Nitazoxanide (1000 mg b.i.d.) combined with the usual dose of atazanavir/ritonavir (300/100 mg od) was well tolerated in this cohort. Laboratory values of haematology and blood chemistry parameters on days 0, 7, and 14 were within normal ranges (Table S3) with no deviations qualifying as grade 1-4 adverse events. In the standard of care plus intervention arm, six patients reported transient known side effects of study drugs (urine discoloration in four and mild abdominal pain in two). No other clinical adverse event was reported.

### Pharmacokinetics of nitazoxanide active metabolite in COVID-19 patients

We compared concentration-time data from day 1 of both periods of the drug-drug interaction study from seven healthy volunteers: 4 females and 3 males aged 24.4 years (4.8) with 56.6 kg (7.5) body weight. Co-administration of nitazoxanide (NTZ) with atazanavir/ritonavir (ATZ/r) increased plasma tizoxanide AUC_0-12_ by from 37.6 µg.h/ml to 63.3 µg.h/ml (68.3%) and its C_max_from 7630 ng/ml to 8730 ng/ml (14.4%) (Figure 5A). A total of 110 concentration-time data were available from the 31 patients in the standard of care plus intervention arm. Median tizoxanide trough plasma concentration was 1546 ng/ml (95% CI: 797-2557), above its putative EC_90_ in 54% of patients^23^ (Figure 5B). An EC_90_ of 1430 ng/ml was reported for nitazoxanide in reversing SARS-CoV-2 induced cytopathic effect in Vero E6 host cells, and tizoxanide is expected to have a similar *in vitro* potency.^9^ Tizoxanide was undetectable in saliva samples collected in the drug-drug interaction study and from patients.

**Figure 5.**
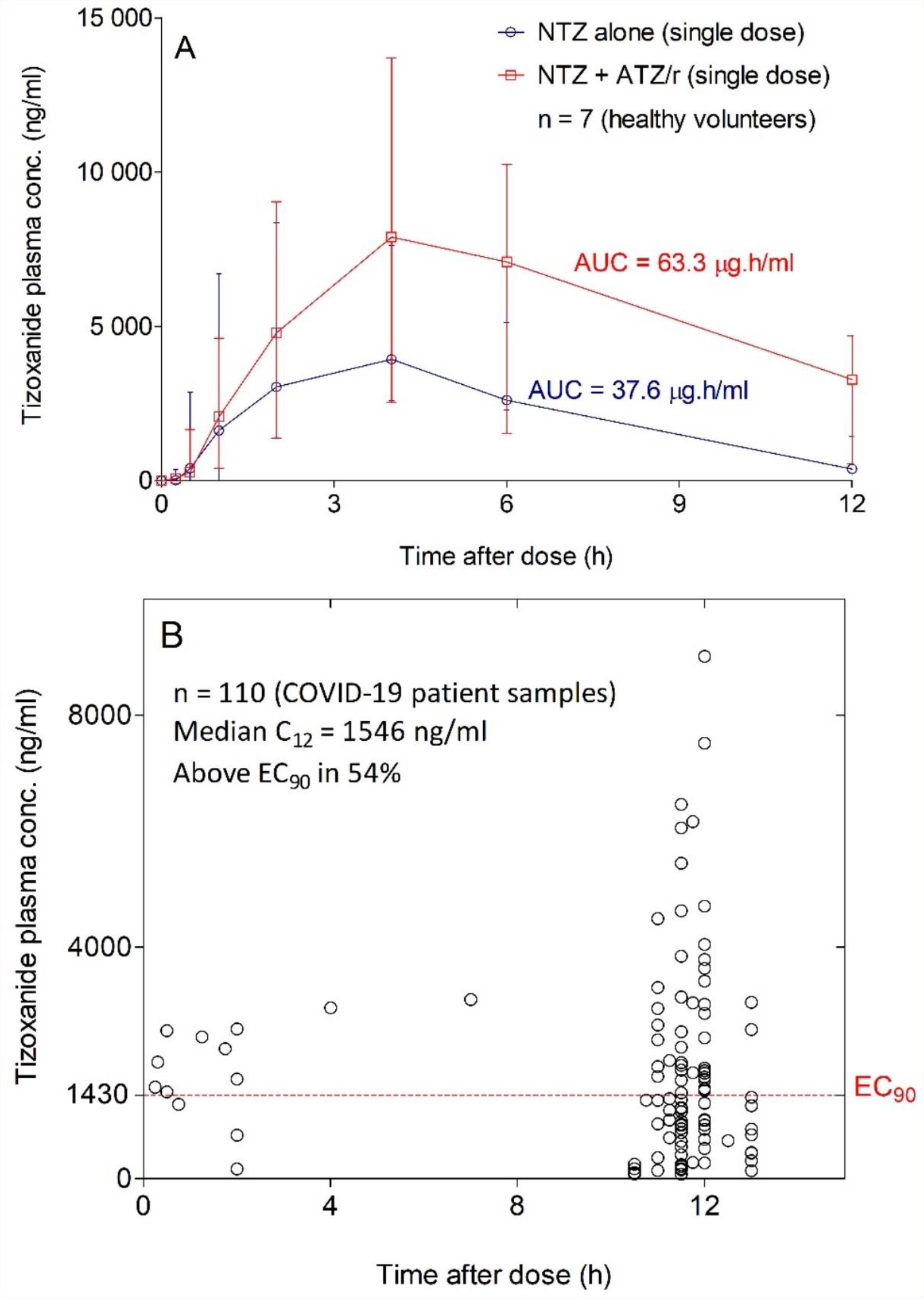
Tizoxanide concentration-time profiles in healthy volunteers and plasma concentration in COVID-19 patients. (A) Co-administration of nitazoxanide (NTZ) with atazanavir/ritonavir (ATZ/r) increased plasma tizoxanide AUC_0-12_ by 68.3% (37.6 µg.h/ml versus 63.3 µg.h/ml) and its C_max_by 14.4% (7630 ng/ml versus 8730 ng/ml). (B) Using samples collected at 11-12 hours after the last nitazoxanide dose (1000 mg b.i.d.), the median concentration was 1546 ng/ml, above the EC_90_ of SARS-CoV-2 in 54% of patient samples.

## Discussion

In this pilot randomised open-label trial, patients who received a 14-day course of nitazoxanide (1000 mg b.i.d.) and atazanavir/ritonavir (300/100 mg od) in addition to standard of care initiated within a few days of COVID-19 diagnosis did not experience a better outcome (clinical improvement, viral clearance, and symptom resolution) compared with those who received standard of care alone. Crucially, tizoxanide plasma exposure was significantly enhanced when combined with atazanavir/ritonavir as expected, possibly *via* inhibition of its inactivation through glucuronidation.^18^ Though concentration in patients at 12 hours after dose was lower than in healthy volunteers, an observation that may be due to the influence of certain components of standard of care, it was above the putative tizoxanide plasma EC_90_ in more than 50% of patients. This is similar to the achievement of plasma concentration above the EC_90_ in 51% of virtual subjects given 1000 mg b.i.d. nitazoxanide with food.^11^ However, tizoxanide was undetectable in saliva samples collected from participants in the drug-drug interaction study and in patients throughout the follow-up period. Tizoxanide is highly bound to plasma proteins (over 99.9%) and we previously highlighted the critical importance of this parameter for *in vitro* to *in vivo* extrapolation.^24^ Our predictions of tizoxanide distribution to human lung^10,11^ based on physicochemical properties, *in vitro* drug binding information, and tissue-specific data did not accurately recapitulate *in vivo* observation.

Confirmation of *in vitro* activity of nitazoxanide against SARS-CoV-2 prompted efforts to investigate its efficacy as a repurposed drug for COVID-19. Several ongoing, completed, or terminated clinical trials include nitazoxanide as monotherapy or as part of a combination strategy. A preprint of interim analysis from a study by Silva *et al* (clinicaltrials.gov identifier: NCT04463264; n = 45) showed no difference in the achievement of PCR negativity by day 7 (62.5% of patients in the 500 mg q.i.d. nitazoxanide arm versus 53.9% in the placebo arm, p = 0.620), though more of those treated with nitazoxanide had viral load reduction of 35% or more from baseline up to day 7 (47.8% versus 15.4%; p = 0.037).^25^ In a preprint of results from the Vanguard study (NCT04486313, n = 379) that enrolled outpatients with mild or moderate COVID-19 within 72 hours of symptom onset, 600 mg b.i.d. extended release nitazoxanide was reported to reduce progression to severe COVID-19 by 85% (1/184, 0.5%) compared with placebo (7/195, 3.6%; p = 0.07). There was no overall difference in time to sustained clinical recovery and a non-significant trend towards a quicker time to symptom resolution and return to usual health was observed.^26^ The Elalfy *et al* study (NCT04392427, n = 113) reported a cumulative day-15 SARS-CoV-2 clearance rate of 88.7% in patients with mild COVID-19 who were treated with a combination of nitazoxanide (500 mg q.i.d.), ribavirin, and ivermectin plus zinc supplement compared with 13.7% in those who received supportive symptomatic therapy (no data on statistical significance).^27^ In the SARITA-2 study (NCT04552483, n = 392), PCR negativity was achieved in 29.9% of patients who received nitazoxanide (500 mg t.i.d. for 5 days) compared with 18.2% in the placebo arm (p = 0.009) and 55% reduction in viral load compared with 45% (p = 0.013). However, there was no difference in symptom resolution between the nitazoxanide and the placebo arms.^28^ Taken together, all three studies where viral load was evaluated reported some benefit, both studies that evaluated symptom resolution observed no benefit, while both studies that evaluated PCR negativity reported conflicting findings.

Similar to the Vanguard and SARITA-2 studies, this analysis of available data from the NACOVID study showed no difference in clinical improvement or symptom resolution between patients treated with standard of care alone versus standard of care plus nitazoxanide (1000 b.i.d.) and atazanavir/ritonavir. However, we only achieved 64% of the target sample size of 89 required to show or exclude 60% improvement in time to SARS-CoV-2 PCR negativity.^20^ Additionally, the limited number of patients with detectable SARS-CoV-2 in saliva at baseline requires that our finding of no difference in viral load change in this trial be interpreted with caution. The choice of saliva for SARS-CoV-2 viral load in this trial was based on observed concordance with nasopharyngeal swabs in the testing laboratory and similar early reports.^29,30^ More recent data now suggest that the suitability of saliva as an alternative to nasopharyngeal swab may be limited to disease stages associated with high viral load.^31,32^ Unfortunately, delays in pre-enrolment testing and diagnosis may have resulted in most patients entering the trial after the exponential phase.

The absence of detectable levels of nitazoxanide active metabolite tizoxanide in saliva samples in this may be indicative of poor penetration into this matrix. If confirmed, this underscores some important points. The use of plasma as a surrogate for target site concentration in COVID-19 should be supported by confirmation of adequate penetration into the respiratory tract and acceptable correlation as with certain antituberculosis drugs.^33^ Remdesivir is known to penetrate poorly into human lungs after intravenous administration,^34^ and nebulised formulation is currently under development^35^ to further enhance its *in vivo* efficacy. Inhalation delivery with targeted activation within the lungs^36,37^ will be an important strategy for drugs with confirmed *in vitro* activity against SARS-CoV-2 but poor penetration into human lungs.

Reports from other completed nitazoxanide studies are pending while several others are still recruiting, including a phase Ib/IIa study investigating within the AGILE clinical trial platform (NCT04746183)^38^ the efficacy of the 1500 mg b.i.d. dosage which was shown to be safe with acceptable tolerability^39^ in mild to moderate COVID-19. As it is unlikely that doses higher than 1500 mg b.i.d. will be tolerable, the AGILE trial is expected to provide a firm signal for whether efficacy can be achieved in COVID-19 at any dose.

## Data Availability

All data produced in the present study are available upon reasonable request to the authors.

## Data sharing statement

The protocol for this clinical trial is already published in BMC Trials and subsequent versions with approved amendments are available upon request. All data collection instruments created for this trial have been made available on REDCap as a project template for other users. Requests for data underlying the outcomes reported in this trial to facilitate further analyses in combination with data from other studies will be considered on a case by case basis. Each request should be accompanied by evidence of ethics approval.

## Acknowledgements

The authors appreciate the patients who participated in this trial at the Infectious Disease Hospital, Olodo, Ibadan, and the Olabisi Onabanjo University Teaching Hospital, Sagamu. We thank the staff and management of both hospitals, as well as Oyo and Ogun State government officials who facilitated the conduct of this trial. We thank Dr Kazeem Akano and Mrs Philomena Eromon who conducted SARS-CoV-2 viral load testing at the African Centre of Excellence in Genomics of Infectious Diseases (ACEGID), Redeemers University, Ede. The Obafemi Awolowo University Bioanalytical Laboratory received infrastructural support from the Liverpool Biomedical Research Centre. The University of Liverpool provided funding for the trial.

## Declaration of interests

We declare no competing interests.

## Contributors

AdO, AnO and SR conceived the initial study. AdO designed the study and developed the protocol with input from all authors. AF, FB, BE, and BOA were responsible for study enrolment and data acquisition. CH was responsible for SARS-CoV-2 viral load determination using RT-PCR. AdO, AA, BA and OOB were responsible for database management and pharmacokinetic analyses. AdO, AA, AF and BE verified the underlying data. AFF and AdO were responsible for analysis and interpretation of data. AdO drafted the manuscript. AnO and OOB critically revised the manuscript. All authors contributed to conducting the trial. All authors revised the report and read and approved the final version before submission. All authors had full access to all the data in the study and had final responsibility for the decision to submit for publication.

## Notes

### Competing Interest Statement

The authors have declared no competing interest.

### Clinical Trial

NCT04459286

### Clinical Protocols

https://trialsjournal.biomedcentral.com/articles/10.1186/s13063-020-04987-8

### Funding Statement

This trial was funded internally by the University of Liverpool and no additional external funding was received.

### Author Declarations

The National Health Research Ethics Committee, Nigeria (approval number: NHREC/01/01/2007-26/08/2020) and the Central University Research Ethics Committee, University of Liverpool (reference number: 8074) approved the protocol. The National Agency for Food and Drug Administration and Control in Nigeria gave regulatory approval for the trial.

