## Supplemental Table 1 for "Efficacy and safety of nitazoxanide combined with ritonavir-boosted atazanavir for the treatment of mild to moderate COVID-19"

**Table S1.** Median time (days) to clinical improvement by participants’ characteristics.

| **Participants** | **All**  **(N = 57)** | **SoC alone**  **(N = 26)** | **SoC with NTZ/ATZ/r**  **(N = 31)** | **Logrank p value** |
| --- | --- | --- | --- | --- |
| **Overall** | 7 | 7 | 8 | 0.913 |
| **Body mass index (kg/m)** |  |  |  | 0.873^a^ |
| Underweight (<18.5) | 9 | 9 | . | 1.000 |
| Normal weight (18.5-24.9) | 7 | 7 | 7 | 0.676 |
| Overweight (24.5-29.9) | 7 | 7 | 7 | 0.352 |
| Obese (≥30) | 9 | 5 | 9 | 0.222 |
| **Age range (y)** |  |  |  | 0.868^a^ |
| 18-50 | 7 | 7 | 8 | 0.921 |
| 51-75 | 7 | 7 | 8 | 0.881 |
| **Sex** |  |  |  | 0.836^a^ |
| Female | 6 | 6 | 6 | 0.793 |
| Male | 8 | 7 | 8 | 0.944 |
| **Comorbidities** |  |  |  | 0.933^a^ |
| No | 8 | 7 | 8 | 0.773 |
| Yes | 7 | 6 | 9 | 0.765 |
| **Time from diagnosis to enrolment (days)** |  |  |  | 0.610^a^ |
| ≤ 1 days | 7 | 7 | 9 | 0.080 |
| 2-4 days | 7 | 7 | 6 | 0.457 |
| ≥ 5 days | 8 | 7 | 8 | 0.411 |
| **Disease severity** |  |  |  | 0.975^a^ |
| Mild Covid-19 | 8 | 7 | 8 | 0.450 |
| Moderate Covid-19 | 7 | 6 | 8 | 0.207 |
| Severe Covid-19 | 4 | . | 4 | 0.157 |
| **Symptoms present at baseline** |  |  |  |  |
| Nose and throat | 7 | 7 | 8 | 0.913 |
| Chest/respiratory | 8 | 6 | 9 | 0.202 |
| Gastrointestinal | 8 | . | 6 | 0.225 |
| Body/systemic | 9 | 6 | 9 | 0.026 |

^a^ Logrank p value for comparison aggregated over stratum-specific results. Abbreviations: SoC, standard of care; NTZ, nitazoxanide; ATZ/r, ritonavir boosted atazanavir.
