## Supplemental Table 2 for "Efficacy and safety of nitazoxanide combined with ritonavir-boosted atazanavir for the treatment of mild to moderate COVID-19"

**Table S2.** Median time to complete symptoms resolution by participants’ characteristics

| **Participants** | **All**  **(N = 57)** | **SoC alone**  **(N = 26)** | **SoC with NTZ/ATZ/r**  **(N = 31)** | **Logrank**  **p value** |
| --- | --- | --- | --- | --- |
| **Overall** | 8 | 6 | 10 | 0.141 |
| **Body mass index (kg/m)** |  |  |  | 0.019^a^ |
| Underweight (<18.5) | 1 | . | . | 0.317 |
| Normal weight (18.5-24.9) | 6 | 2 | 9 | 0.022 |
| Overweight (24.5-29.9) | 9 | 7 | 10 | 0.222 |
| Obese (>=30) | 10 | 6 | 13 | 0.348 |
| **Age and age range (y)** |  |  |  | 0.234 ^a^ |
| 18-50 | 8 | 4 | 11 | 0.083 |
| 51-75 | 9 | 10 | 6 | 0.716 |
| **Sex** |  |  |  | 0.156 ^a^ |
| Female | 9 | 5 | 10 | 0.450 |
| Male | 7 | 6 | 9 | 0.231 |
| **Comorbidities** |  |  |  | 0.199 ^a^ |
| No | 8 | 4 | 10 | 0.122 |
| Yes | 7 | 7 | 9 | 0.982 |
| **Time from diagnosis to enrolment (days)** |  |  |  | 0.044 ^a^ |
| ≤ 1 days | 5 | 6 | 6 | 0.293 |
| 2-4 days | 10 | 7 | 11 | 0.271 |
| ≥ 5 days | 5 | 1 | 6 | 0.032 |
| **Disease severity** |  |  |  | 0.084 ^a^ |
| Mild Covid-19 | 6 | 4 | 9 | 0.230 |
| Moderate Covid-19 | 10 | 10 | 14 | 0.262 |
| Severe Covid-19 | 6 | . | . | 0.317 |
| **Symptoms present at baseline** |  |  |  |  |
| Nose and throat | 8 | 6 | 10 | 0.141 |
| Chest/respiratory | 10 | 7 | 10 | 0.687 |
| Gastrointestinal | 10 | . | 10 | 0.479 |
| Body/systemic | 10 | 10 | 5 | 0.963 |

^a^ Logrank p value for comparison aggregated over stratum-specific results. Abbreviations: SoC, standard of care; NTZ, nitazoxanide; ATZ/r, ritonavir boosted atazanavir.
