## Supplemental Table 3 for "Efficacy and safety of nitazoxanide combined with ritonavir-boosted atazanavir for the treatment of mild to moderate COVID-19"

**Table S3.** Mean (SD) value of haematological and blood chemistry parameters (safety tests) on days 0, 7 and 14.

| **Safety test** | **Day 0 (n = 44)** | | **Day 7 (n = 44)** | | **Day 14 (n = 44)** | |
| --- | --- | --- | --- | --- | --- | --- |
|  | **SoC** | **SoC + NTZ plus ATZ/r** | **SoC** | **SoC + NTZ plus ATZ/r** | **SoC** | **SoC + NTZ plus ATZ/r** |
| **Haematology** |  |  |  |  |  |  |
| White blood cells count (x10^6^/L) | 5805 (2301) | 4941 (1090) | 6026 (2529) | 5614 (1247) | 6057 (1702) | 5755 (1397) |
| Red blood cells count (x10^6^/µL) | 5.21 (0.61) | 5.36 (0.54) | 5.01 (1.50) | 5.31 (0.53) | 5.16 (0.64) | 5.25 (0.56) |
| Haemoglobin (g/dL) | 13.3 (1.60) | 14.0 (1.66) | 12.9 (3.84) | 14.3 (0.83) | 13.7 (1.43) | 13.4 (2.76) |
| Haematocrit (%) | 39.8 (4.82) | 42.2 (4.75) | 38.5 (11.5) | 41.5 (7.44) | 41.0 (4.60) | 42.0 (2.90) |
| Mean corpuscular volume (fL) | 78.2 (5.50) | 80.4 (6.48) | 74.5 (21.8) | 81.8 (7.15) | 78.5 (5.11) | 80.6 (6.40) |
| MCH (pg) | 24.3 (3.22) | 24.6 (3.31) | 24.4 (7.21) | 25.2 (3.64) | 25.6 (2.58) | 25.4 (3.80) |
| Platelets (x10^9^/L) | 196 (93.4) | 205 (57.4) | 235 (118) | 211 (51.3) | 229 (105) | 236 (52.3) |
| Neutrophils (%) | 45.8 (13.2) | 38.3 (11.1) | 45.0 (16.1) | 41.7 (10.9) | 47.1 (12.0) | 41.5 (10.4) |
| Lymphocytes (%) | 42.4 (12.7) | 49.1 (10.6) | 42.1 (15.0) | 46.9 (10.2) | 42.8 (11.3) | 47.3 (9.95) |
| Eosinophils (%) | 2.80 (2.66) | 3.20 (2.40) | 2.42 (1.65) | 3.68 (3.39) | 2.58 (1.97) | 3.59 (2.65) |
| Basophils (%) | 0.23 (0.46) | 0.14 (0.09) | 0.17 (0.12) | 0.14 (0.11) | 0.22 (0.16) | 0.17 (0.25) |
| Reticulocyte | 4.55 (1.99) | 7.04 (4.99) | 4.68 (2.20) | 6.98 (7.50) | 3.60 (1.43) | 4.21 (2.54) |
| **Blood chemistry** |  |  |  |  |  |  |
| Alanine aminotransferase (IU/L) | 29.0 (17.0) | 27.7 (9.26) | 31.3 (14.3) | 33.9 (14.5) | 37.0 (12.5) | 33.5 (12.1) |
| Aspartate aminotransferase (IU/L) | 34.4 (13.2) | 34.6 (12.2) | 34.7 (17.7) | 38.8 (15.6) | 65.1 (43.8) | 45.0 (20.1) |
| Bilirubin (mg/dL) | 0.51 (0.73) | 0.59 (0.50) | 0.72 (0.80) | 1.75 (2.10) | 0.91 (1.47) | 1.96 (1.62) |
| Alkaline Phosphatase (IU/L) | 57.1 (17.9) | 62.2 (16.5) | 57.3 (21.4) | 63.1 (18.7) | 66.0 (15.8) | 70.2 (20.1) |
| Albumin (g/dL) | 4.42 (0.84) | 4.67 (0.42) | 4.20 (1.22) | 4.58 (0.49) | 4.68 (0.46) | 4.68 (0.50) |
| Total protein (g/dL) | 7.26 (1.72) | 7.84 (0.62) | 7.21 (2.07) | 7.86 (0.55) | 7.49 (0.52) | 7.71 (0.89) |
| Sodium (mEq/L) | 136 (4.22) | 138 (4.71) | 131 (38.4) | 137 (3.21) | 137 (2.06) | 136 (2.78) |
| Potassium (mEq/L) | 3.79 (0.42) | 4.26 (0.80) | 3.71 (1.10) | 4.09 (0.52) | 3.83 (0.44) | 3.84 (0.36) |
| Chloride (mEq/L) | 103 (10.4) | 102 (4.34) | 95.3 (27.8) | 102 (5.70) | 99.8 (3.19) | 101 (2.87) |
| Bicarbonate (mEq/L) | 23.7 (3.25) | 23.3 (2.96) | 22.6 (6.63) | 25.3 (2.40) | 23.3 (1.52) | 24.7 (3.73) |
| Blood urea nitrogen (mg/dL) | 22.2 (9.99) | 22.9 (8.21) | 24.4 (11.1) | 21.2 (6.97) | 25.9 (9.75) | 23.1 (10.6) |
| Creatinine (mg/dL) | 0.96 (0.24) | 0.99 (0.29) | 0.90 (0.28) | 1.15 (1.21) | 0.93 (0.16) | 0.95 (0.26) |

Abbreviations: SoC, standard of care; NTZ, nitazoxanide; ATZ/r, ritonavir boosted atazanavir.
